# The effect of emotion regulation on emotional eating among undergraduate students in China: the chain mediating role of impulsivity and depressive symptoms

**DOI:** 10.1101/2023.01.09.23284356

**Authors:** Huimin Yang, Xinyi Zhou, Longjiao Xie, Jing Sun

## Abstract

This study aimed to examine the relationship between difficulties in emotion regulation and emotional eating and the role of impulsivity and depressive symptoms in mediating this chain. Four hundred ninety-four undergraduate students participated in the study. A self-designed questionnaire was used in the survey from February 6 to 13, 2022, to finish our purpose, including the Emotional Eating Scale (EES-R), Depression Scale (CES-D), the Short Version of the Impulsivity Behavior Scale (UPPS-P) and Difficulties in Emotion Regulation Scale (DERS). Results showed that 1) There was a positive correlation between difficulties in emotion regulation, impulsivity, depressive symptoms, and emotional eating, respectively.2) Impulsivity and depressive symptoms mediated the relationship between emotion regulation and emotional eating separately. 3) Impulsivity and depressive symptoms played the mediating chain role between emotion regulation and emotional eating.

## Introduction

Eating disorders are characterized by persistent disordered eating behaviors that interfere with daily social and psychological functioning[1]. The global incidence of eating disorders in 2019 was 41.9 million all over the world and caused 6.6 million Disability-Adjusted Life Years (DALYs) in that year [2]. Patients with eating disorders described their binge eating behaviors as a result of negative emotions[3]. Emotional eating (EE) is combating negative emotions by engaging in binge eating behaviors[4, 5]. The number of people who have EE was continually increasing over the past years [6]. EE affects people of all ages, and approximately 20% of people regularly adopt emotional eating behaviors[7]. The researcher found that adults aged 21 to 39 are much more likely to have EE [7]. Ashley’s study noted that approximately 10% to 60% of adolescents are emotional eaters[8].

Evidence of a follow-up study showed that EE impel people to overeat and cause weight gain[9]. Notably, EE was particularly related to the preference for sweet and high-fat foods,which can lead to excessive intake of high energy-density foods and promote the occurrence of obesity[10]. Researchers also pay attention to the relationship between EE and weight loss[11], and found that EE hinder the weight loss[12], and the higher level of EE related to the less weight loss over the same period[13, 14]. As an abnormal eating behavior, EE is the trigger for gastrointestinal disorders, which was found to be a factor causing pharyngeal reflux and acid reflux[6].

Emotion regulation is defined as the ability to cope with negative emotions in an adaptive way[15]. Difficulties in emotion regulation were found among obese adolescence with emotional eating[16], which was proven to be one factor of overeating[17, 18]. Poor emotion regulation was related to impulsivity[19] and depression [20], both of which were showed have a positive connection with emotional eating[21, 22].

Impulsivity is recognized as the rapid and unplanned reactions to stimulation without thinking about the negative consequences of these reactions[23]. It is a multidimensional construct that involves attentional, behavioral, cognitive components [24] and has been reported frequently for years[25, 26]. Data from a large national sample of the United States population showed that nearly 20% of the participants have impulsivity, especially among younger individuals [27]. 40% of children and 58.3% of adults had higher clinically elevated impulsivity indicated by the data in 2011[28].

Depression is an unusually low and unpleasant mood-altering negative emotional state that harms personal life and society [29].The World Health Organization reports that the global prevalence of depression is as high as 12.8%[30]. College students have more negative emotions due to their high academic pressure, uncertain employment prospects and other problems[31]. Study elucidated that the prevalence of depressive symptoms among college students is increasing[32].Approximately 23.8% of first-year university students have depressive symptoms[33] and more than 30% of university students have depression[34].

Although emotional eating has been studied for years, few studies have examined emotional eating among undergraduate students and the specific mechanism between emotion regulation and emotional eating. Thus, in this study, we discuss the effect of emotion regulation on emotional eating and the internal mechanisms among undergraduate students, that is, the mediating role of impulsivity and depressive symptoms in the relationship between emotion regulation and emotional eating. The study aim to give scientific suggestions to reduce the incidence of emotional eating. The results stress the significance of regulating emotions in a correct and efficient way, while paying attention to the impulsivity and depressive symptoms among undergraduate student and giving an early intervention also can contributes to solve the problem.

### Emotion regulation and emotional eating

Difficulties in emotion regulation lead to unhealthy eating behaviors[35, 36], especially binge eating[37]. The escape model explained that individuals who have difficulties in emotion regulation may erode the usual inhibitions around food, and made people be willing to escape the negative emotions by breaking their dietary rules and restricting their focus to eating itself[38, 39]. Emotion regulation and eating disorders had positive relations, proofed by a multiple correlation analysis in Italian university students[40]. Gianini investigated 326 obese and binge-eating adults and found that difficulties in emotion regulation can predict emotional eating[37]. A study among 552 undergraduate students demonstrated that difficulties in emotion regulation contribute greatly to emotional eating[36]. Although previous studies have proven the relationship between emotional regulation and emotional eating, the relationship still needs to be explored in Chinese undergraduate students. Therefore, we put forward the following hypothesis:

H1: Difficulties in emotion regulation are positively correlated with emotional eating among Chinese undergraduate students.

### The mediating role of impulsivity

Impulsivity is one of the mental dimensions taking part in emotion regulation[41], which is generally regarded as a consequence of impaired executive functioning. Prior work proved that emotion dysregulation was related to impulsive behaviors(r=0.31,p< 0.1)[42]. Multivariable analysis indicated that eating disorders are positively associated with impulsivity in obese people[43]. A sample contained 121 obese participants with binge eating behaviors were proved having high impulsivity with average score of 31.11 assessed by UPPS-P Impulsive Behavior Scale[43].In conclusion, emotional regulation may influence emotional eating through impulsivity. Therefore, we put forward the following hypothesis:

H2: Impulsivity plays a mediating role in Difficulties in Emotion Regulation and Emotional Eating among Chinese undergraduate students.

### The mediating role of depressive symptoms

Depression was taken for a result of inappropriate emotion regulation[44].A study conducted among university students indicated that, individuals with depression had more difficulties in emotion regulation[45]. The results from random effects analyses pointed those difficulties in emotion regulation was connected with depressive symptoms[46]. Researchers used an emotion-provoking film to increase the negative emotion of participants and showed that difficulties in emotion regulation were related to higher levels of depression symptoms [47]. Braden carried out an experiment among adults with overweight or obesity and indicated that taking eating as a response to depression was most closely related to emotion regulation difficulties [48]. A study conducted in a random sample of 10,000 people in Finland revealed that emotional eating is associated with an increase of depression[49].To summarize, difficulties in emotion regulation related to depressive symptoms, and depressive symptoms can predict emotional eating. Thus, we pose Hypothesis 3:

H3: Depressive symptoms play a mediating role in Difficulties in Emotion Regulation and Emotional Eating among Chinese undergraduate students.

### The chain mediating role of emotion regulation and emotional eating

Increased impulsivity and negative mood may be amplified by difficulties in emotion regulation among overweight people, which was found by Leehr’s experiment using electroencephalography and eye tracking[50]. A study among adolescents aged 13–21 years implied that difficulties with impulse control can result in emotional eating when they have a negative mood[51].A large cohort of study conducted inn adults revealed that, impulsivity, as a distinct personality factor, gives rise to one set of depressive illnesses in adults [52]. Previous research in adolescents have suggested that impulsivity is related to rumination, self-blaming and catastrophe[53]. Impulsivity also lead adolescents to encounter adverse situations, which, in turn, result in depression[54]. Above all, difficulties in emotion regulation issue in emotional eating and are also related to impulsivity and negative mood. Meanwhile, depression is a typical negative mood. Therefore, we speculate that impulsivity and depressive symptoms can mediate the relationship between emotion regulation and emotional eating. Therefore, we have the following hypothesis:

H4: Impulsivity and depressive symptoms play a chain mediating role in difficulties in emotion regulation and emotional eating among Chinese undergraduate students.

## Materials and methods

### Data source

According to the principle of multivariate statistical analysis, the estimated sample size was 5 times the number of observation indexes[55]. Given that there is a potential 10 percent missing samples, 467 samples were planned to be included. A questionnaire survey was developed and administered from February 6 to 13, 2022. This study selected students who were in a university. The exclusion criteria were ① completing the questionnaire in less than 100 seconds, ② the choices showed a clear pattern, and ③ age was older than 18 years or younger than 26 years. By using a professional platform named Wenjuanxing (HTTP://www.xjn.cn), the questionnaires were distributed to the students via We Chat, which is an app that supplies communication services and is used widely in China. Students who saw the questionnaires and were willing to participate could complete them via mobile phone during free time. The registered users monitored the collected questionnaires in no time and eliminated invalid questionnaires with no self-reported hearing impairment, with all the same answers, or with inconsistent forward and reverse answers.

Research Ethics Committee approval was obtained from the IRB of Peking University (No. IRB00001052-21150). Before conducting the online survey, the participants were informed that involvement was completely voluntary and anonymous. In addition, we introduced the purpose of the study again in the guidelines of each questionnaire and emphasized the confidentiality of the survey.

### Measures

#### Emotional eating

Zhu Hong (2012) revised the Chinese version of the Emotional Eating Scale (EES-R), adding the positive emotional dimension to the original scale[56]. There are 23 items divided into four dimensions: depression, anxiety, anger and positivity. Responses ranged from 1 to 5, with 1 representing a total lack of these emotions and 5 indicating a high degree of emotion. The higher the score, the higher the desire to eat in a certain mood. The Cronbach’s alpha coefficient in this study was 0.935.

#### Emotion regulation

Gratz developed the difficulties in Emotion Regulation Scale (DERS) and his team in 2004[57]. The scale is a total of 36 items, which can be divided into six dimensions: difficulty in emotional awareness, difficulty in emotional understanding, difficulty in impulse control, difficulty in goal orientation, difficulty in accepting emotional responses, and difficulty in effectively using regulatory strategies. Each item’s responses range from 1 (never) to 4 (always). Item 11 is scored in reverse. The higher the score is, the more serious the emotion regulation difficulty is and the lower the emotion regulation ability is. In this study, the Cronbach’s alpha coefficient was 0.934.

#### Impulsivity

The short version of the UPPS-P Impulsive Behavior Scale (UPPS-P) is a 20-point evaluation of five different impulsive personality traits that demonstrate conceptual validity [58]. Responses to questions ranged from 1 (strongly agree) to 4 (strongly disagree), with higher scores indicating stronger negative urgency. In this study, the Cronbach’s alpha coefficient was 0.834.

#### Depressive symptoms

The Center for Epidemiological Survey, Depression Scale (CES-D) was developed by Radloff, National Institute of Psychiatry in 1977[59]. The scale has a total of 20 items, each of which measures one symptom. According to the frequency of symptoms occurring in the last week, each item response ranged from 0 (none or very few) to 3 (disagree strongly), and 4, 8, 12 and 16 were scored in reverse. A score less than or equal to 15 indicates no depressive symptoms, 16-19 is likely to have depressive symptoms, and 20 indicates depression symptoms. The Cronbach’s alpha coefficient was 0.884.

### Data analysis

All statistical analyses were performed with SPSS 25.0. At the outset of the data analysis, descriptive statistics and statistical analyses were used to assess whether emotional feeding was correlated with other variables in the intended direction. The Pearson product-moment correlation coefficient (r) was used in the correlation analysis. The range from -1 to 1 and the values of 0.1, 0.3, and 0.5 represent “small”, “medium”, and “high” correlations, respectively [60]. In the hierarchical regression analysis, the contribution of all these components to emotional eating was examined by including all components of reading fluency in the final model. Model 6 of the SPSS process plug-in was used to test the chain mediation model. For the significance test of the regression coefficient, the bootstrapping method with 5,000 repeated samples was selected to obtain a robust[61].

## Results

### Sociodemographic characteristics and descriptive analysis

A total of 506 questionnaires were collected across 25 Chinese provinces (N= 264 in Beijing, N=80 in Guangdong, N= 19 in Shandong and N= 131 in other provinces). The final sample size was 494, with an effective valid response rate of 97.7%.

The sample of this study were undergraduate students whose average age was 20.1 ± 1.9 years old. The descriptive statistics are shown in Table 1. The majority of the participants was female (62.8%), while 184 males were included. There were 340 junior grade students in our study, accounted for 68.8%, and the rest of participants were senior students (N=154,31.2%).

**Table 1.** Descriptive statistics of demographic variables (N=494)

| Characteristics | N (%)<br>/Mean (SD) | Emotional<br>Eating median<br>(IQR) | r/F | P |
| --- | --- | --- | --- | --- |
| Age | 20.13(1.858) |  | 0.220*** | 0.000 |
| BMI | 21.64(5.111) |  | 0.006 | 0.887 |
| Gender |  |  | 3.427** | 0.001 |
| Male | 184(37.2) | 64.00(24.00) |  |  |
| Female | 310(62.8) | 58.50(21.00) |  |  |
| Grade level |  |  | 0.193 | 0.661 |
| Junior | 340 (68.8) | 59.55(22.00) |  |  |
| Senior | 154 (31.2) | 64.42(23.00) |  |  |
| Average<br>Monthly<br>Income |  |  | 4.552* | 0.011 |
| ¥0-3000 | 60(12.1) | 69.00(25.50) |  |  |
| ¥3000-5000 | 165(33.4) | 60.00(21.00) |  |  |
| ¥5000 or more | 269(54.5) | 60.00(24.00) |  |  |

Note: \*p<0.05, \*\*p<0.01, \*\*\*p<0.001

Emotional eating was associated with age, gender, grade and income. Statistical analyses indicated that age (r = 0.220; P = 0.000) and gender (t = 3.427; P = 0.001) were significantly associated with emotional eating. Males were reported greater emotional eating than females. Senior grade in university reported insignificantly greater emotional eating than junior (P = 0.661). Individuals with a monthly income less than ¥3000 reported significantly greater emotional eating compared to individuals making more than ¥3000 a month (P = 0.011).

### Pearson correlation analysis

There was a significantly correlated relationship among difficulties in emotion regulation, impulsivity, depressive symptoms and emotional eating. As shown in Table 2, Pearson correlation analyses revealed that there was a medium positive correlation of emotional eating and depressive symptoms (r= 0.363, p<0.01), difficulties in emotion regulation (r= 0.260, p<0.01) and impulsivity (r= 0.382, p<0.01). Impulsivity was found to be moderately positive correlated with depressive symptoms (r= 0.422, p<0.01) and difficulties in emotion regulation (r= 0.473, p<0.01). The correlation between depressive symptoms and difficulties in emotion regulation was high (r= 0.656, p<0.01). The results showed that there were significant positive correlations between difficulties in emotion regulation, impulsivity and depressive symptoms, supporting hypothesis 1.

**Table 2.** Descriptive statistics and correlations among study variables (N=494)

|  | M | SD | 1 | 2 | 3 | 4 |
| --- | --- | --- | --- | --- | --- | --- |
| 1 CES-D | 19.83 | 12.094 | 1.00 |  |  |  |
| 2 DERS | 101.27 | 18.810 | 0.656*** | 1.00 |  |  |
| 3 S-UPPS-P | 51.73 | 8.021 | 0.422*** | 0.473*** | 1.00 |  |
| 4 EES | 61.06 | 17.460 | 0.363*** | 0.260*** | 0.382*** | 1.00 |

Note: \*\*\* $p < 0.001$

**Table 3.** Hierarchical regression analysis with emotion regulation, impulsivity, depression and emotional eating.

| Variables | Model 1 |  |  | Model 2 |  |  | Model 3 |  |  | Model 4 |  |  |
| --- | --- | --- | --- | --- | --- | --- | --- | --- | --- | --- | --- | --- |
|  | EES-R |  |  | UPPS-P |  |  | CES-D |  |  | EES-R |  |  |
| | $\beta$ | t | P | $\beta$ | t | P | $\beta$ | t | P | $\beta$ | t | P |
| DERS | 0.221 | 5.098*<br>** | 0.000 | 0.445 | 11.067<br>*** | 0.000 | 0.572 | 15.042<br>*** | 0.000 | -0.060 | -1.083 | 0.279 |
| UPPS-P |  |  |  |  |  |  | 0.127 | 3.314*<br>* | 0.001 | 0.283 | 6.093*<br>** | 0.000 |
| CES-D |  |  |  |  |  |  |  |  |  | 0.247 | 4.548*<br>** | 0.000 |
| R2 | 0.121 |  |  | 0.246 |  |  | 0.462 |  |  | 0.228 |  |  |
| F | 13.464<br>*** |  | 0.000 | 31.796<br>*** |  | 0.000 | 69.824<br>*** |  | 0.000 | 20.555<br>*** |  | 0.000 |

Note: \* $p < 0.05$ , \*\* $p < 0.01$ , \*\*\* $p < 0.001$ ; DERS, difficulties in emotion regulation; UPPS-P, impulsivity; CES-D, depression; EES-R, emotional eating

### Hierarchical regression analysis

Impulsivity may mediate difficulties in emotion regulation and emotional eating. At the first stage (Model 1), difficulties in emotion regulation were added to the model and significantly predicted emotional eating (β = 0.221, p < 0.001). At the second stage (Model 2), impulsivity was included in the model, and difficulties in emotion regulation significantly predicted impulsivity (β = 0.445, p < 0.001). At the third stage (Model 3), difficulties in emotion regulation (β = 0.572, p < 0.001) and impulsivity (β = 0.127, p < 0.05) significantly and positively predicted depressive symptoms. In the fourth stage (Model 4), the addition of impulsivity and depressive symptoms to the model led to a distinct change in the beta value of difficulties in emotion regulation, and difficulties in emotion regulation no longer made a significant contribution to emotional eating. These results indicate that impulsivity and depressive symptoms may have a chain mediating role in the relationship between difficulties in emotion regulation and emotional eating.

### Mediation analysis

The PROCESS macro of SPSS was suitable for the analysis of the effects of mediation and for chain mediation models with multiple mediating variables[62]. Therefore, we used Model 6 in the PROCESS macro of SPSS to analyze the chain mediation effect of impulsivity and depressive symptoms between difficulties in emotion regulation and emotional eating. Taking emotion regulation difficulties as the independent variable, emotional eating as the dependent variable, and impulsivity and depressive symptoms as the mediating variable, the chain mediation effect test yielded the results shown in Fig 1 and Table 4. Our study examined the gender, age, grade and income mediating effects as control variables. The results show that difficulties in emotion regulation have a significant indirect effect on emotional eating through impulsivity (β= 0.1172, 95% CI= [0.07,0.17]); that is, impulsivity has a significant mediating effect on the relationship between emotional regulation difficulties and emotional eating. Thus, difficulties in emotion regulation have significant indirect effects on emotional eating through depressive symptoms (β= 0.1316, 95% CI= [0.07, 0.20]); that is, depressive symptoms have a significant mediating effect on the relationship between difficulties in emotion regulation and emotional eating. Finally, the indirect effect of difficulties in emotion regulation on emotional eating through impulsivity and depressive symptoms was significant (β= 0.0130, 95% CI= [0.00, 0.03]). In other words, impulsivity can affect depressive symptoms. At the same time, the chain mediation path from difficulties in emotion regulation to impulsivity, then to depressive symptoms, and finally to emotional eating is significant, supporting Hypothesis 2. The results indicate that impulsivity and depressive symptoms play a continuous mediating role in the relationship between difficulties in emotion regulation and emotional eating.

**Fig 1.**
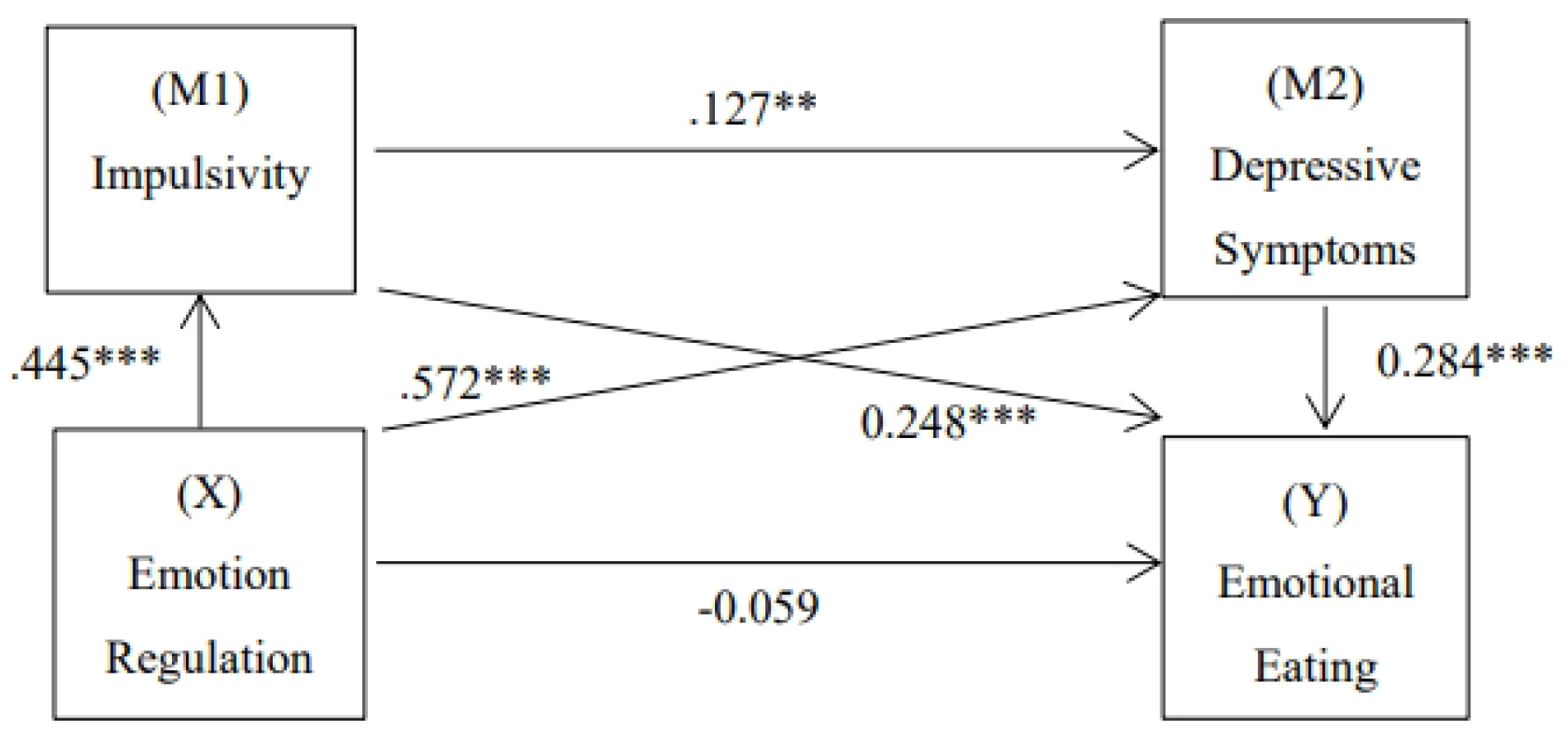
The role of impulsivity and depressive symptoms as chain mediators in the relationship between emotion regulation difficulties and emotional eating with standardized beta. Note: ***p<0.001. Values on paths are path coefficients (standardized βs).

**Table 4.**
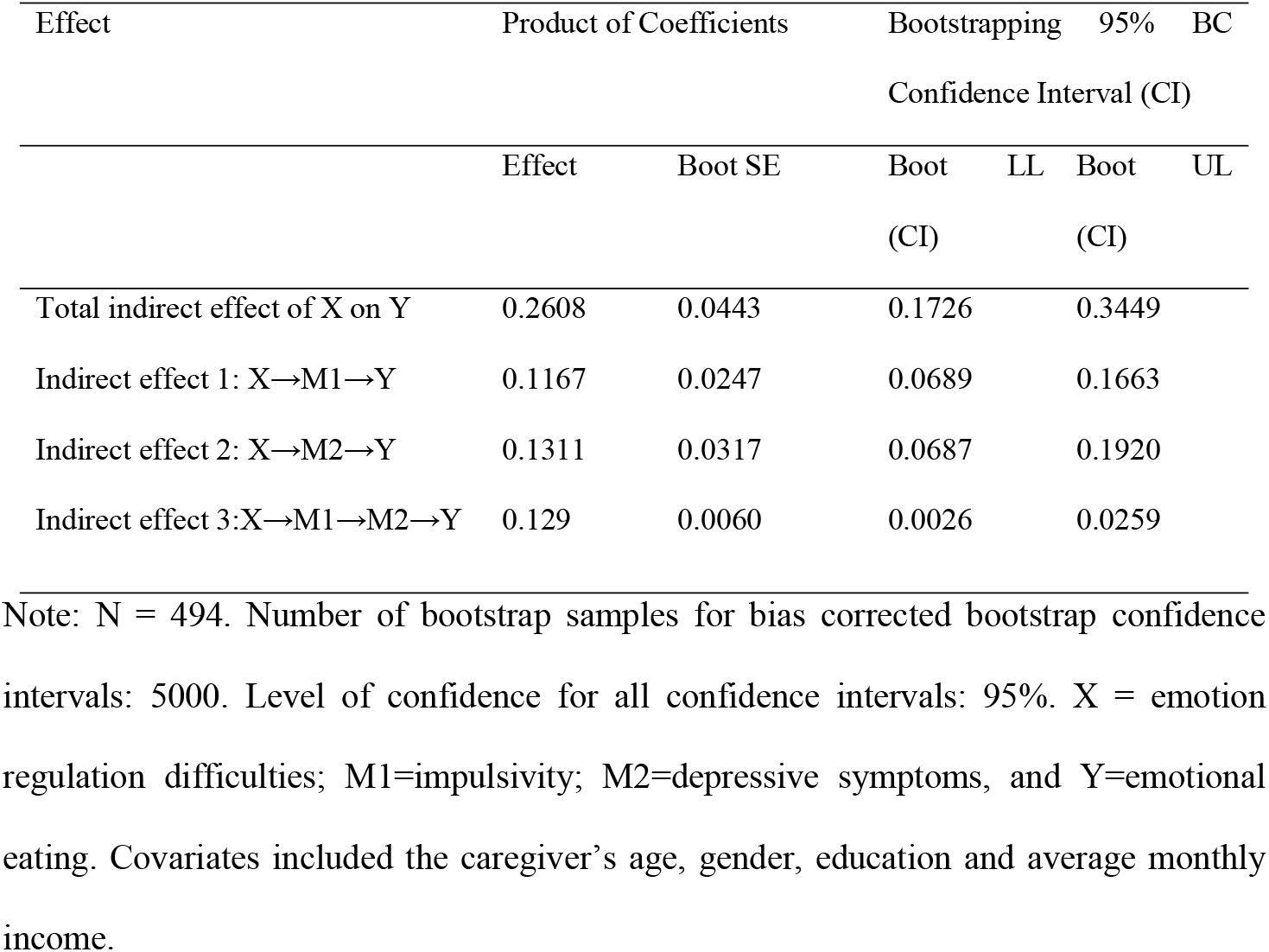
Chain-Mediated Effects of Difficulties in Emotion Regulation on Emotional Eating Through Impulsivity and Depressive Symptoms.

## Discussion

### The status of emotional eating

In our study, the average score of emotional eating among undergraduate students was 61, higher than the score 52 in Shi’s study[63]. Our results showed that the senior students were more likely to engage in emotional eating. It can be explained that, the senior students usually face more employment and academic pressure as they are near the graduation[64]. The current data pointed out that male have a higher level of emotional eating than female, which is contrary to some previous studies[6, 65]. It can be explained that women are more concerned with weight control and body shape maintenance[66]. Average monthly income is also a factor that influences emotional eating in our study, the lower income indicates higher level of emotional eating, which is consistent with a previous report[67]. Hemmingsson’s theory emphasized that low income may cause negative mood, which finally induce eating energy-dense foods to alleviate negative emotions and stress [68]. Burdick proved that lower income is associated with greater impulsivity[69]. While people with higher impulsivity were influenced deeply by negative emotions, which was appeared as the cause of emotional eating[70].

### Difficulties in emotion regulation are positively correlated with emotional eating

Based on our study, difficulties in emotion regulation had a positive predictive effect on emotional eating, which verifies H1. People who have difficulties in emotion regulation are more likely rapped in negative emotions[71]. A review reported that people with depression use less emotion regulation strategies[72]. People with difficulties in emotion regulation cannot understand the feelings they have, and this lack of emotional clarity make for eating disorder[73]. Difficulties in emotion regulation lead an inability to deal with tasks[74] and to use effective regulatory strategies[57], after then increase the probability for eating as an attempt to escape from the averse emotional states[48]. Under the negative circumstances, people choose eating as a strategy because they can feel happy in this way [75], moreover, eating more calories in the first 5 minutes can make them more positive[76]. Dopamine is a probable explanation; eating increases dopamine release by glutamatergic projection to the ventral tegmental area mediates and NMDA receptor, which then causes the feeling of pleasure to people.[77, 78]. Above all, emotion regulation showed a positive connection with emotional eating.

### The relationship between difficulties in emotion regulation and emotional eating is mediated by impulsivity

The current study found that impulsivity played a mediating role between difficulties in emotion regulation and emotional eating, which verifies H2. Valente (2017) suggested that emotional eating is an iceberg top of a three-step ladder and that the hidden base of the iceberg is represented by both emotional dysregulation and the level of impulsivity [79]. Difficulties in emotion regulation especially impulse control may decrease DAD_2_ receptor availability in the striatum and result in lower DA-argic activity, which signified a tendency of impulsivity[80]. Bratec (2017) found that difficulties in emotion regulation reduce the influence of Prefrontal Cortex on ventral striatal aPE signals after then change the normal status of striatum[81]. Individuals who cannot activate striatal circuits appropriately, perhaps have impaired self-regulatory control, which contributes to impulsive behaviors[82]. Overeating behavior has been confirmed to have association with impulsivity[83]. Impulsivity may contribute to overeating in situations of uncontrolled emotion[84]. Thus, difficulties in emotion regulation can induce emotional eating through impulsivity.

### The relationship between difficulties in emotion regulation and emotional eating is mediated by depressive symptoms

The current study found that depressive symptoms played a mediating role between difficulties in emotion regulation and emotional eating, consistent with previous studies [85], which verifies H3. Martin et al proved the difficulties in emotion regulation may be a predictor of depression in general adults[86]. Difficulties in emotion regulation would lead to added negative moods and might result in depressive symptoms after some time [87, 88]. Longitudinal research found that eating can be a way to temporarily numb uncomfortable emotions, such as depression. Individuals who experiencing depression may use food as entertainment[89]. Eating triggered by depression was closely associated with emotion regulation difficulties[48]. It is possible that when in negative emotions (e.g., depression), individuals would increase eating behavior as a strategy for regulating. The 5-HTTLPR is proved to be associated with depression[90]. Additionally, a study proved that adolescents with depressive symptoms showed more increase in emotional eating if they carried the 5-HTTLPR genotype[91]. Thus, difficulties in emotion regulation can induce emotional eating through depression.

### Impulsivity and depressive symptoms are chain mediators in the relationship between difficulties in emotion regulation and emotional eating

Our results showed the chain mediation of difficulties in emotion regulation on emotional eating through impulsivity and depressive symptoms, which verifies H4. The results of our research is consistent with previous study, which proved that impulsivity can predict the onset of depression as a distinct personality factor in adults by logistic regression model[54]. Individuals who have difficulties in emotion regulation are at significant risk of using inappropriate strategies to cope with life events[92]. Past research found that the higher emotion dysregulation group scored significantly higher on impulsivity[93]. Low levels of 5-HT and 5-HIAA were found in people classified as impulsive[94-96] and levels of 5-HT and 5-HIAA have positive relationship with severity of depression and could be good markers for evaluating depression[97]. Our results suggested that difficulties in emotion regulation may have a tendency toward impulsivity and depression. By effectively acting as mediators through impulsivity and depressive symptoms, people might have emotional eating when have difficulties in emotion regulation[98]. The current study is the first to document a sequential process in which difficulties in emotion regulation affect impulsivity, which in turn predicts depression and thus over eating.

## Limitation

One limitation of the study is that the results cannot be generalized to overall population, because undergraduate students differ in social pressure and environment from others. The sample was underrepresented to demonstrate the overall situation of college students in China because the sample size is limited and sample sources cannot cover all regions of China. Emotional eating, emotion regulation, impulsivity and depressive symptoms were measured by self-report, the results are not as accurate as laboratory studies or observational measures in daily routine. We can’t make sure that each participant understands the question well and if they wrote a wrong answer which was Different from the reality. The participants may have false memories or lie to us.

A laboratory studies or observational measures is needed to assess emotional eating, difficulties in emotion regulation, impulsivity and depressive symptoms to further explore this research question. Future studies should investigate the different results of emotional eating after intervening in the two mediation variables in the current study to explore the impact on emotional overeating.

## Conclusion

Through our study, it can be seen that the state of emotional eating among Chinese university students is grim. The status of emotional eating seems to be associated with difficulties in emotion regulation. It is time to explore the relationship because emotional eating can increase the risk of obesity and other diseases. The relationship between difficulties in emotion regulation and emotional eating is mediated by impulsivity and depression symptoms, respectively. In addition, impulsivity and depressive symptoms are chain mediators in the relationship between the two. The current study provides data support for improving the status of emotional eating and the reference direction for the formulation of interventions.

## Data Availability

All relevant data are within the manuscript and its Supporting Information files.

## Acknowledgement

We wish to thank all the college students who take part in our questionnaire investigation. Funding from the Peking University Students Innovation Project is also gratefully acknowledged.

